# 3D-Printed Prosthetic Socket Adapter for Clinical Simulation Use in Resource-Limited Environments

**DOI:** 10.1101/2025.08.03.25332912

**Authors:** Prince Oduro, Cecil Owusu Bempah, Sadat Osei-Wusu, Joel Adjei Boateng, Daniel Opoku-Gyamfi, Kannan Govindan, Napoleon Abiwu

## Abstract

**Background:** Prosthetics and Orthotics (P&O) education in low-income nations faces significant challenges due to the prohibitive cost and limited availability of essential training materials, particularly prosthetic components like socket adapters. This scarcity directly impedes practical skill development, forcing a reliance on theoretical instruction.

**Methodology:** A structured methodology integrating CAD modeling (Siemens NX12), Finite Element Analysis (FEA) using Siemens NX Nastran, and Fused Deposition Modeling (FDM) with PLA filament on a Creality Ender 3 printer was employed. FEA simulations guided design refinements.

**Findings:** Physical prototype testing confirmed outstanding results. The adapter demonstrated dimensional accuracy with less than 0.1 mm deviation using a digital Vernier caliper, crucial for interoperability. Mechanical compression testing on an Instron 5944 UTM showed the prototype withstood a maximum load of 600 N before significant deformation, far exceeding simulation needs. Furthermore, fitting and compatibility tests confirmed a seamless and secure fit with standard transtibial prosthetic components. The final print quality was visually satisfactory with minimal post-processing.

**Conclusion:** This work validates 3D printing as a highly effective, affordable, and reproducible solution for creating functional P&O training tools. Our findings offer a significant step towards democratizing access to practical prosthetic education globally, establishing a replicable framework for developing essential, locally manufactured training aids in various engineering and medical fields.

## Introduction

The design of prostheses, with its origins tracing back to ancient Egyptian and Roman civilizations, has continuously evolved to restore functionality and facilitate societal reintegration for individuals with limb loss [1]. However, the global landscape of Prosthetics and Orthotics (P&O) education faces a significant, often overlooked, challenge: the prohibitive cost and limited availability of teaching and training materials, particularly for essential prosthetic components needed for clinical simulation practice [2–4]. This issue is felt in lower-income nations, where, according to the World Health Organization, only 5% to 15% of individuals requiring prostheses can access them due to material scarcity and high prices [5, 6]. This scarcity extends critically into the educational sphere, directly impacting the quality and practical depth of P&O training programs [7–9]. While the emergence of 3D printing offers a promising avenue for cost-effective and personalized component fabrication, its potential to address the pedagogical gap in P&O education, specifically regarding the provision of affordable training tools, remains underexplored.

Despite the availability of computer-aided design (CAD) and computer-aided manufacturing (CAM) techniques for prostheses since the 1980s, their widespread adoption, especially in educational settings, has been gradual [10–12]. Transtibial (below-knee) amputation is the most common type of limb loss, and individuals with such amputations rely on prostheses comprising elements like the socket, socket adapter, pylon, suspension mechanism, and foot [13, 14]. A fundamental aspect of P&O education involves understanding the biomechanical interactions between the residual limb, the prosthetic socket, and other components like the socket adapter, which are vital for optimizing device performance [2, 15].

The core problem addressed by this work stems directly from the critical lack of affordable and accessible teaching materials for P&O education and training, particularly for components like prosthetic socket adapters in low-resource setting. In contexts such as Ghana, the availability of these essential teaching aids is severely restricted [16]. This limitation is primarily driven by the high cost of importing prosthetic components, inflated by taxes and unfavorable currency exchange rates, coupled with significant shipping delays [4, 17]. The consequence is that prosthetic socket adapters become largely inaccessible, not only for practicing prosthetists but, more critically, for prosthetics trainees and students who need hands-on experience during their studies [16]. Due to unavailability of locally made affordable components for practical and clinical simulation learning, P&O education often defaults to theoretical instruction, hindering the development of crucial fabrication and fitting skills. Furthermore, traditional fabrication methods including paper drawing designs, casting, CNC machining, and manual welding are inherently time-consuming, labor-intensive, and require expensive machinery and materials [18–20]. This makes them unsuitable for rapid, cost-effective replication needed for extensive student training [20]. In contrast, 3D printing offers rapid, on-demand part production with minimal setup, at a significantly lower cost (a 3D printer and its components can be up to ten times cheaper than CNC machining equipment) and requires substantially less workshop space [21, 22].

To bridge this critical gap in P&O education and training, this project introduces an innovative solution: the design and fabrication of a prototype prosthetic socket adapter using a 3D plastic filament. This work directly aims to enhance the pedagogical resources available for prosthetics students in low-resource setting by leveraging the advantages of 3D printing, including rapid prototyping for iterative design improvements without expensive tooling, and inherent cost-effectiveness for producing customized components. This approach promises to democratize access to practical training materials, allowing students to engage in hands-on learning with realistic, functional prototypes that are otherwise prohibitively expensive or unavailable. Ultimately, this research seeks to revolutionize the teaching, learning, and practical training experiences for aspiring prosthetists in Ghana and similar low-resource settings.

## Methodology

The development of this 3D-printed prosthetic socket adapter was guided by a structured methodology that integrated design, computational analysis, and fabrication into a single, cohesive process. The overarching goal was to create a low-cost, reproducible, and effective training tool for Prosthetics and Orthotics (P&O) students in resource-limited settings. The project began with a needs assessment, which identified a critical gap in educational resources: the high cost and limited availability of standard prosthetic components for hands-on clinical simulation instruction. This directly informed the project’s primary objectives: to produce a functional prototype that was both affordable and easy to reproduce using widely available Fused Deposition Modeling (FDM) technology.

### Design and Material Selection

The first critical step was the material selection, as the choice of 3D-printing filament would directly influence the cost, mechanical performance, and accessibility of the final product. After a comprehensive review of common thermoplastics including Acrylonitrile Butadiene Styrene (ABS), Polyethylene Terephthalate Glycol (PETG), Thermoplastic Polyurethane (TPU), and Polyether Ether Ketone (PEEK), Polylactic Acid (PLA) was chosen as the most suitable material. This decision was based on a careful balance of factors: PLA offers an exceptional combination of low cost and ease of printing, which are paramount for educational initiatives in resource-constrained environments. While its limitations regarding moisture sensitivity and a relatively low glass transition temperature were acknowledged, these were deemed acceptable for a proof-of-concept prototype intended for educational simulation demonstrations rather than long-term clinical use.

With the material determined, the CAD modeling phase commenced using Siemens NX12. The adapter’s geometry was designed to be compatible with common industry standards for transtibial socket attachments. The male pyramid adapter is the type of adapter designed for this project. The design process was iterative, focusing on integrating essential features such as standardized fastening holes for universal component attachment, textured surfaces for grip, and reinforced structural elements to distribute stress efficiently. Detailed 2D sketches illustrating the key dimensions of the adapter design are presented in Figure 1, providing a comprehensive overview of its geometry and critical interfaces. These conceptual drawings were then translated into a precise 3D digital model within Siemens NX12, as shown in Figure 2, which served as the foundation for subsequent analysis and 3D printing.

**Figure 1:**
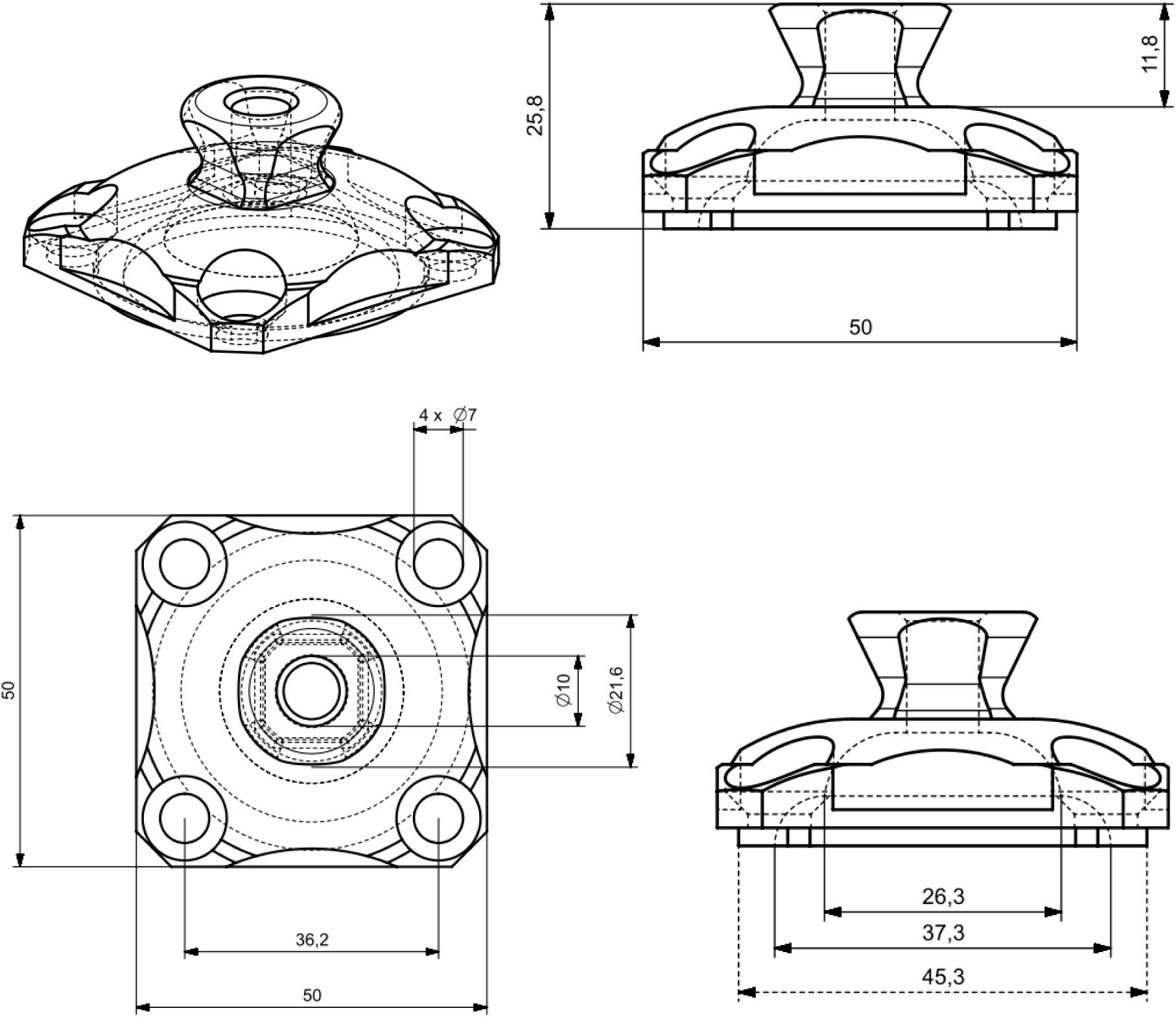
2D Sketches with Dimensions of the socket adapter.

**Figure 2:**
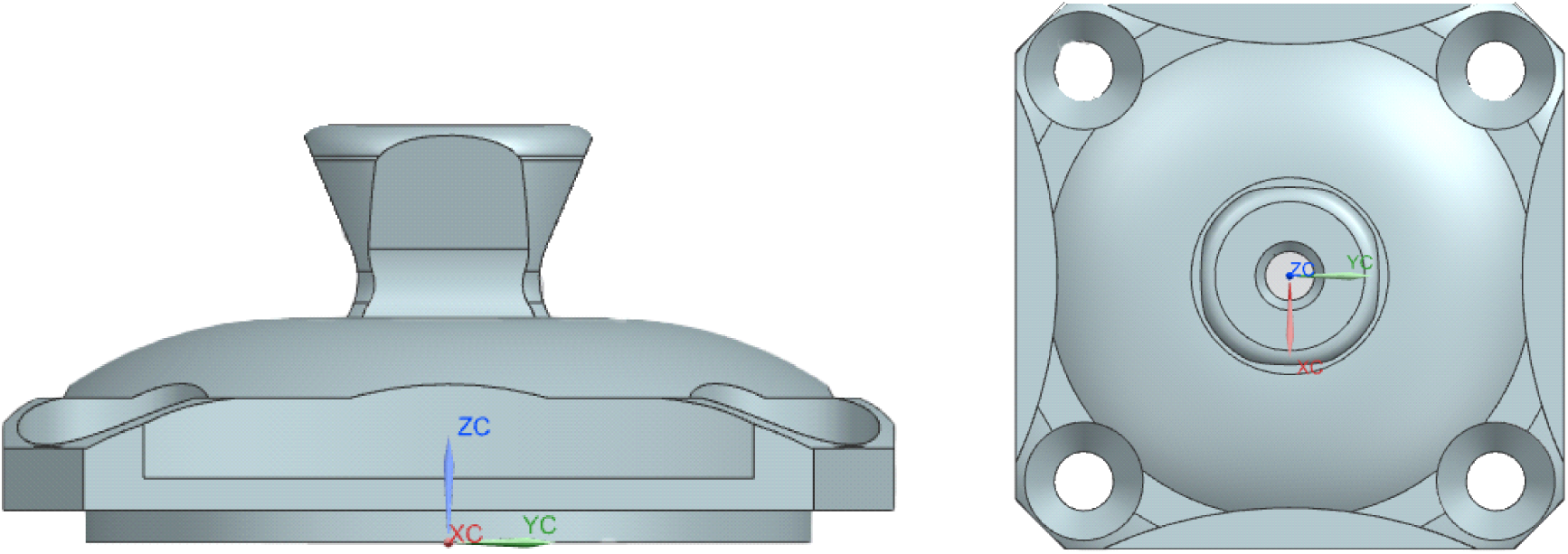
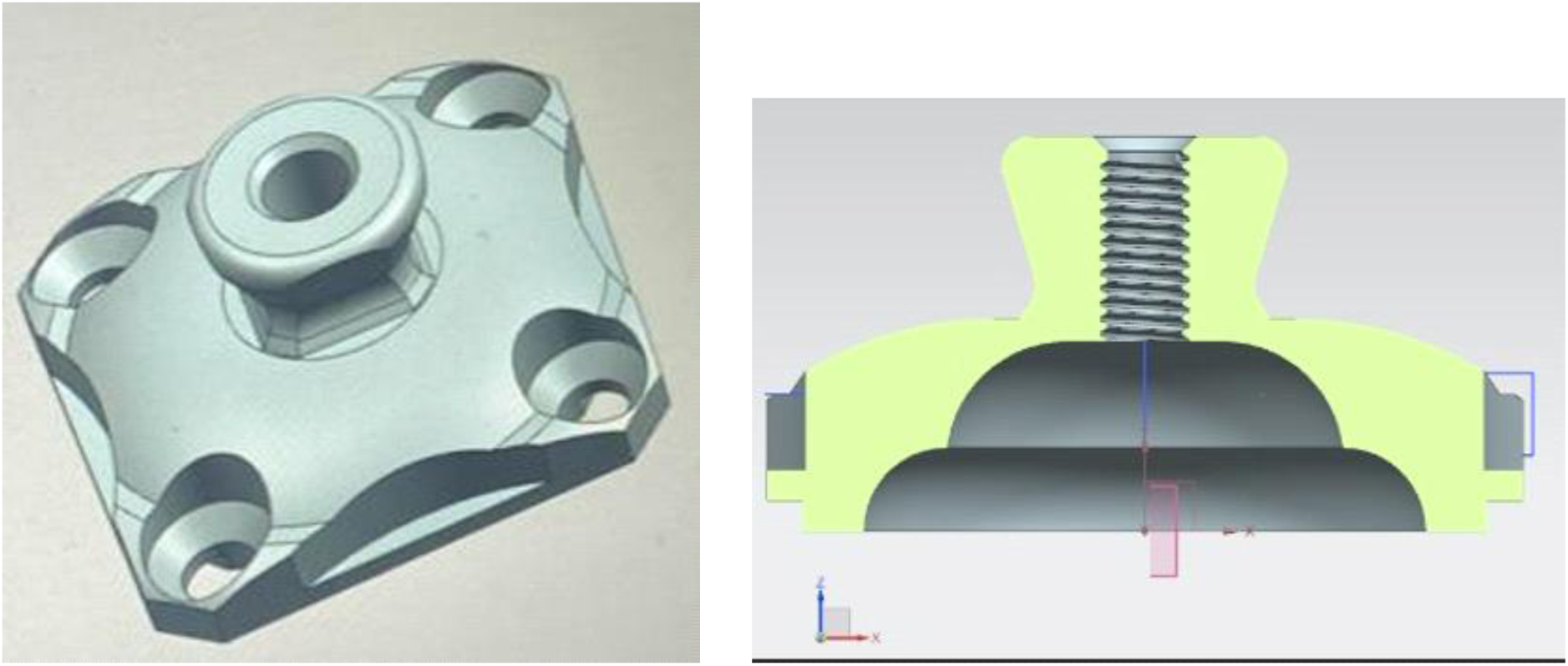
3D Models of the pyramid.

To ensure the design’s structural integrity, a Finite Element Analysis (FEA) was performed using Siemens NX Nastran. The simulation was configured to replicate typical in-use loading conditions. Specifically, a static load of 500 N (axial compression) and 200 N (lateral shear) was applied, representing a simulated peak forces experienced by a prosthetic component during the stance phase of gait. The FEA results identified critical stress concentrations, particularly at the interface between the adapter and the pyramid receiver. These findings directly informed subsequent design modifications, leading to localized thickening of the walls and the addition of internal rib reinforcements. The final design was then assessed for theoretical compliance with the structural integrity requirements outlined in ISO 10328:2016 before being exported as a stereolithography (STL) file.

### 3D Printing and Post-Processing

The final prototype was produced using a Creality Ender 3 FDM printer and blue PLA filament. Prior to printing, the machine underwent a thorough calibration process to ensure optimal performance. The nozzle temperature was set to 220°C and the heated build plate to 60°C to prevent warping and promote strong first-layer adhesion. The STL file was then prepared using the Ultimaker Cura slicer software, a crucial step that generated the G-code, the specific set of instructions understood by the 3D printer for executing the print as shown in figure 3.

**Figure 3:**
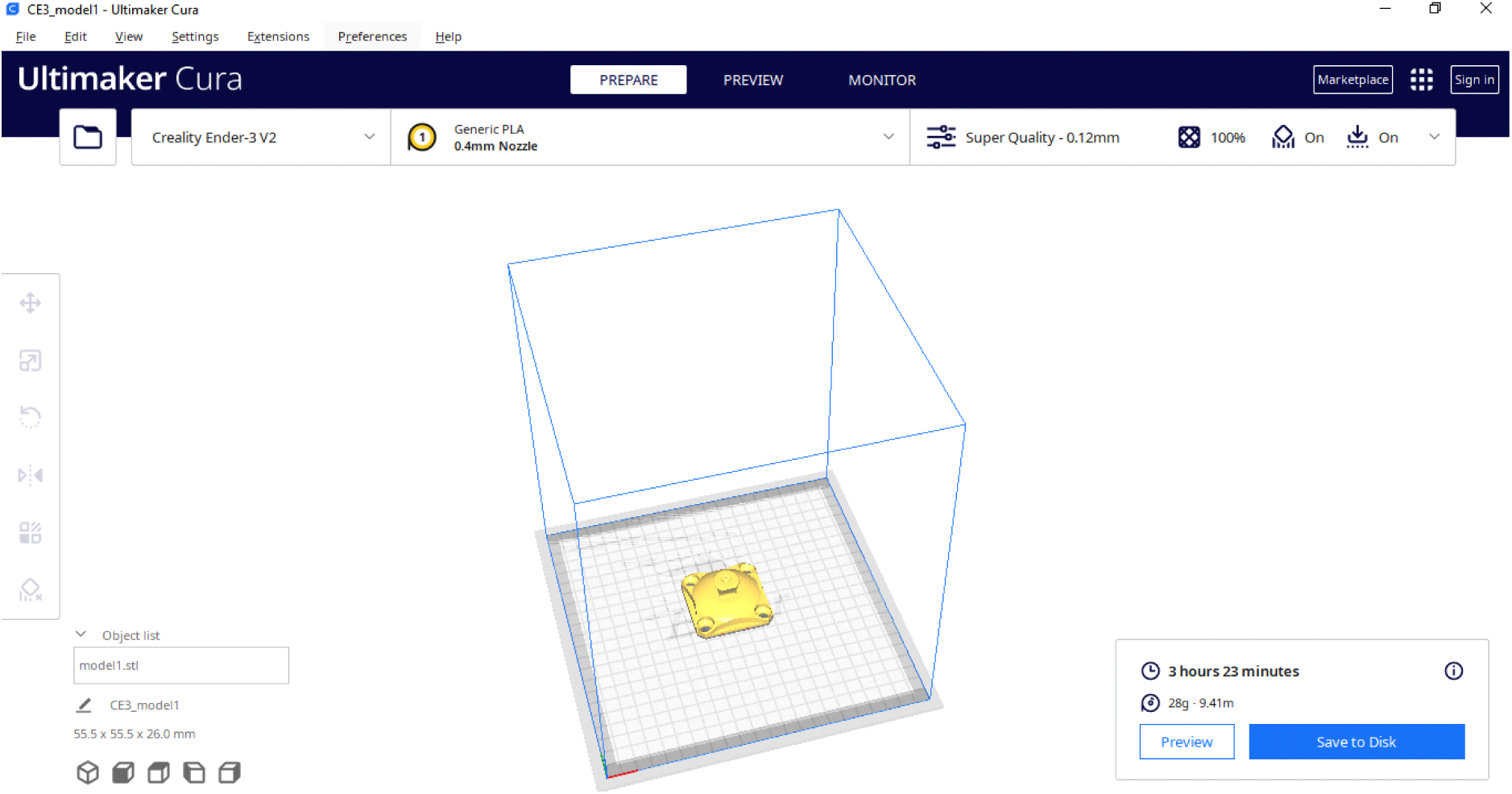
Ultimaker Cura slicing interface.

During the slicing process, key parameters were meticulously configured to maximize the prototype’s strength and precision. Each adapter required approximately 28g of PLA filament. The estimated time for printing generated by Cura the software used for slicing was 3 hours 23 minutes. A fine layer height of 0.12 mm was chosen to achieve optimal print quality and a smooth surface finish, crucial for representing intricate details accurately. Based on the structural requirements and the desire for maximum mechanical strength for educational demonstration, the infill density was set to 100% with a cubic pattern. The print speed was adjusted to 700 mm/s to ensure efficient fabrication. Furthermore, support structures were enabled and automatically generated by the slicer to manage overhangs and complex geometries effectively. To ensure robust bed adhesion and prevent warping, a brim was opted for, providing an enlarged contact area during printing. Throughout the print, the first layer was closely monitored, with minor manual adjustments made as necessary.

## Results

This chapter presents the comprehensive findings derived from the computational analysis and the subsequent physical testing of the 3D-printed prosthetic socket adapter prototype. These results meticulously detail the prototype’s performance under simulated and real-world conditions, validating its technical viability in the context of its design specifications.

### Computational Analysis Results

The Finite Element Analysis (FEA) provided an indispensable insight into the prototype’s structural behavior under the prescribed simulated load conditions, critically informing the final design iterations. The displacement analysis, visually represented through color-coded maps, graphically depicted the component’s deformation profile. This analysis revealed a range of displacements, from negligible values (approaching 0.000 mm, depicted in dark blue) to a significant maximum of 0.136 mm, observed specifically at the adapter’s central feature, particularly within its neck region. The full extent of this deformation is illustrated in Figure 4, where areas in red and orange clearly indicate regions of highest displacement. This critical finding directly informed a crucial design iteration: the neck region, identified as a point of excessive deformation in initial simulations, was subsequently strengthened and thickened to enhance its structural integrity (as conceptually depicted in the design evolution illustrated in Figure 5).

**Figure 4:**
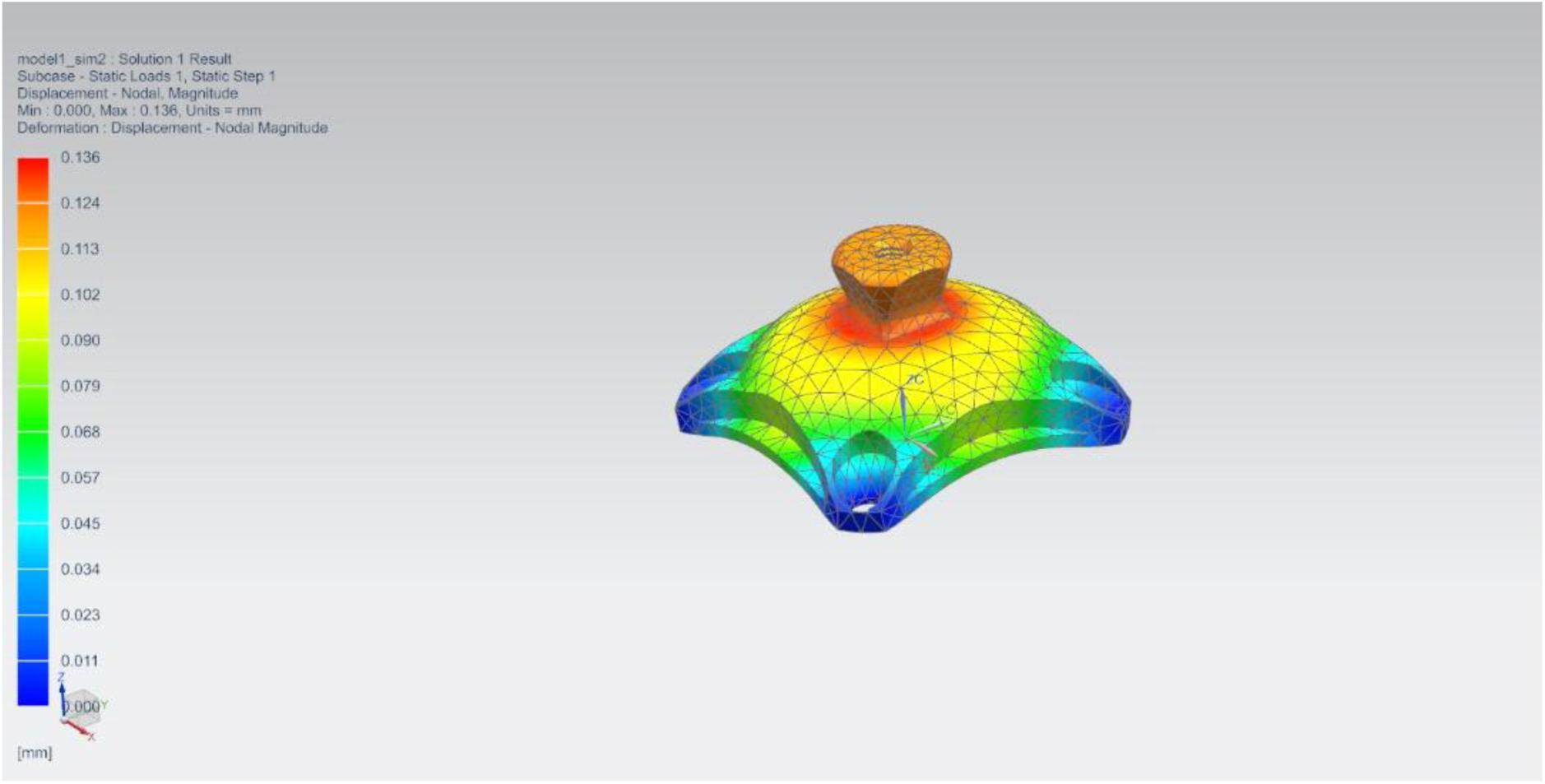
Finite element analysis results interface

**Figure 5:**
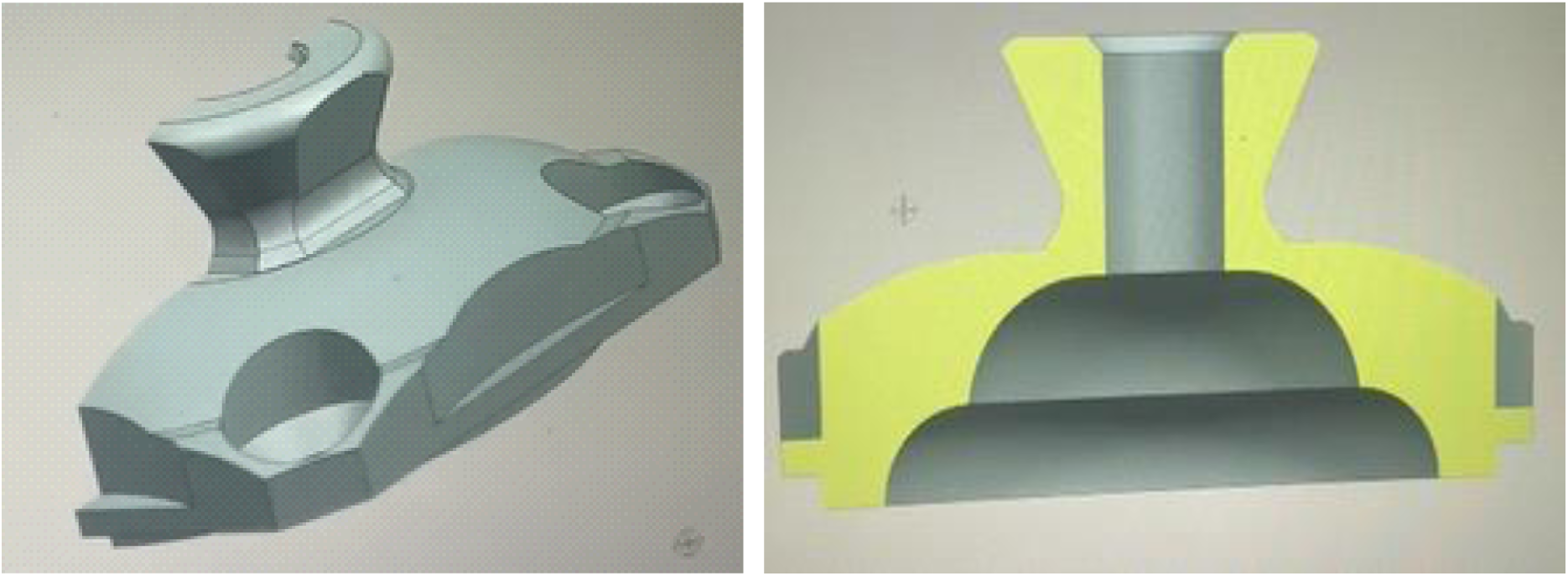
New design made with strengthened neck region after the FEA demonstrated highest deformation at the neck region.

Following the displacement analysis, a detailed FEA for Von Mises stress distribution was performed. This analysis indicated stress levels ranging from a minimal 0.02 MPa (blue) across less loaded areas to a peak of 64.81 MPa (red). High-stress concentrations were predominantly observed around the central feature and in localized areas near the screw holes, confirming these as critical load-bearing regions. The maximum stress of 64.81 MPa was precisely located at the junction of the central feature with the main adapter body. This uneven load distribution, keenly highlighted by the analysis, reinforced the necessity of the aforementioned design modifications, aiming to achieve a more balanced stress profile and mitigate potential material failure under expected prosthetic loads comprehensively visualized in Figure 6.

**Figure 6:**
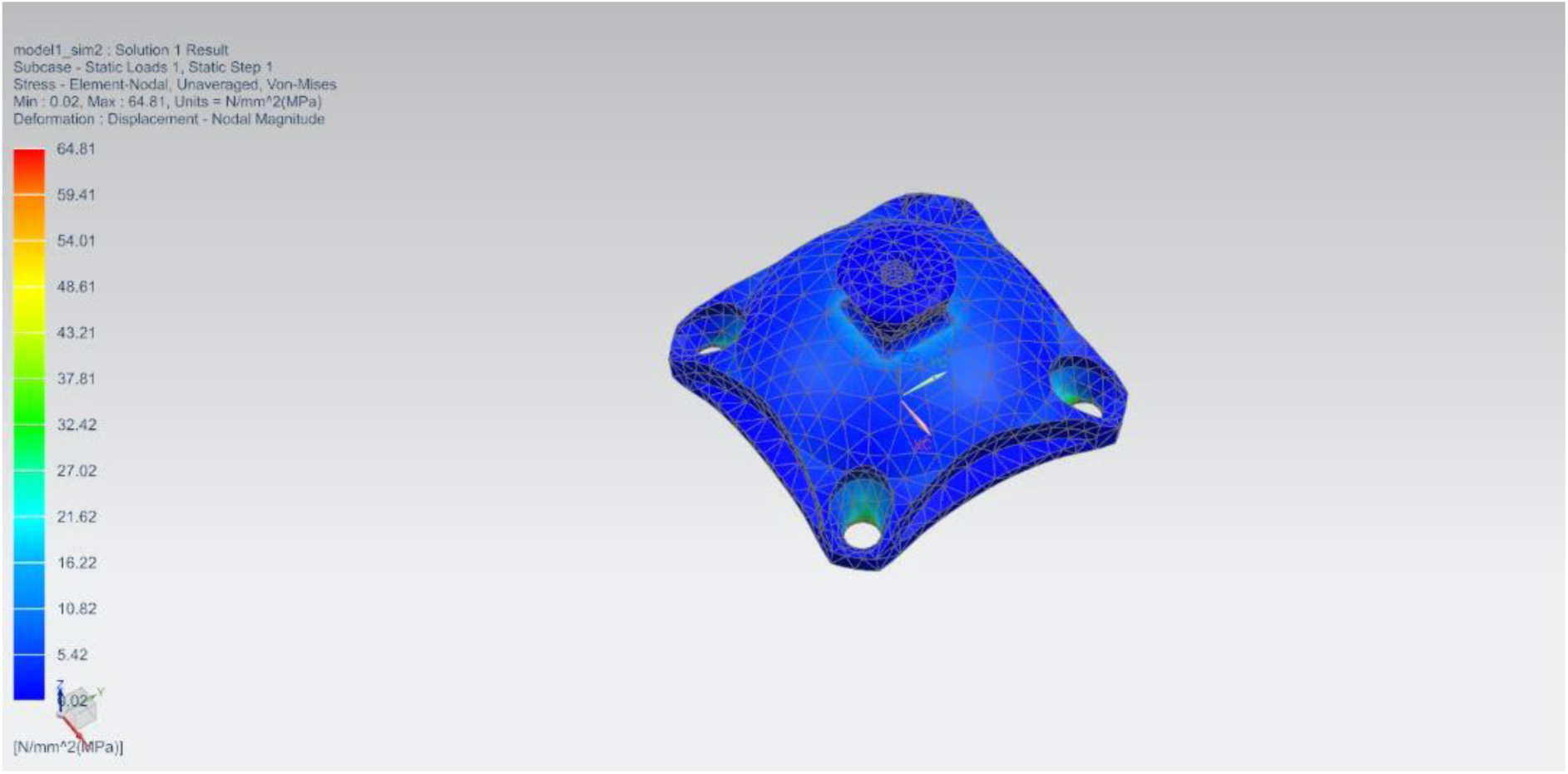
Stress distribution analysis

### Post-printing

The entire 3D printing process took approximately 3 hours, 25 minutes. Upon completion, the printed adapter was allowed to cool completely before being carefully removed. This involved the meticulous removal of support structures, light sanding to smooth rough edges, and a final inspection. A surface finish and quality inspection were carried out to evaluate the aesthetic and tactile attributes of the prototype. This involved a visual examination of the prototype to identify any surface defects, prominent layer lines, or irregularities caused by the 3D printing process. A tactile examination was also performed to assess surface smoothness. The surface finish was generally found to be smooth, with minimal visible layer lines, indicating good print quality. Although some localized areas required minor post-processing to achieve a finer finish, the overall quality of the prototype was deemed satisfactory for a prototype at this stage and suitable for its role as a simulation educational tool. Figure 7 provides a visual representation of the final 3D-printed prosthetic socket adapter, showcasing its completed form and surface quality.

**Figure 7:**
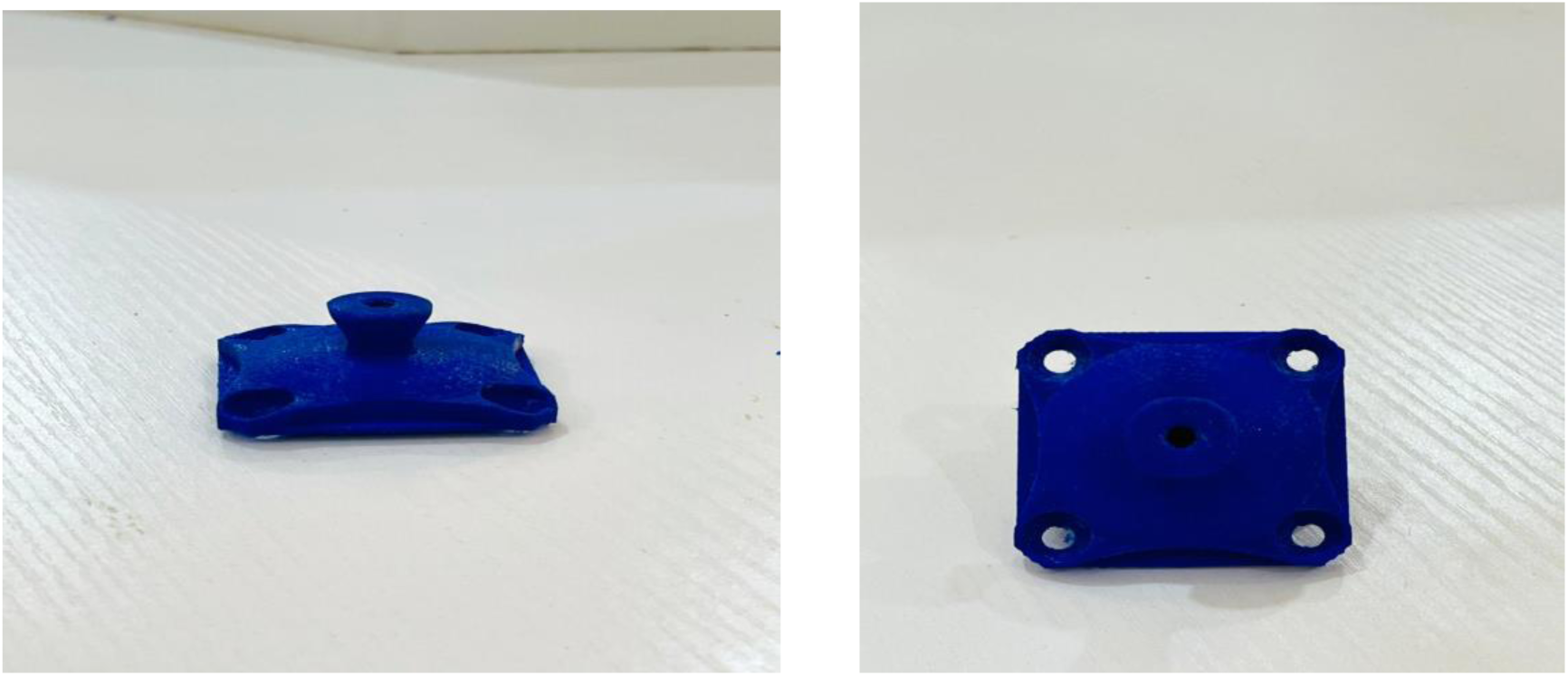
3D printed prosthetic socket adapter.

### Prototype Testing and Evaluation

Following the computational analysis, a comprehensive series of physical tests was performed on the successfully 3D-printed prototype to thoroughly assess its dimensional accuracy, mechanical integrity, fitting, and overall quality. This rigorous evaluation phase was key in determining whether the prototype met the established design specifications and functional requirements for simulation educational applications.

A rigorous dimensional accuracy test was first conducted to ensure the prototype’s dimensions accurately matched the specifications outlined in the CAD model. Critical dimensions of the fabricated adapter were measured using a digital Vernier caliper, focusing on areas such as the diameter of the screw holes, the height of the central feature, and the overall width and length of the adapter base. The measurements consistently confirmed that the prototype was well within the acceptable tolerance range, with deviations of less than 0.1 mm in most dimensions. This validated that the 3D printing process maintained a high level of dimensional accuracy, crucial for component interoperability.

For mechanical testing, the objective was to determine the load-bearing capacity of the prototype under simulated real-world conditions. A compression test was performed using an Instron 5944 universal testing machine (UTM), where incremental loads were applied to the adapter. The load was gradually increased until the component exhibited significant deformation. The prototype successfully withstood a maximum load of 600 N before showing signs of significant plastic deformation. This result was well within the expected range for its intended low-stress, clinical simulation-based application, indicating the adapter’s capability to support the necessary forces in a practical training scenario.

A crucial fitting and compatibility test was then conducted to assess that the prototype fits correctly with the corresponding components of a standard prosthetic system. The procedure involved assembling the 3D-printed adapter with a standard transtibial prosthetic limb setup, checking for proper alignment, secure fit, and ease of assembly. The prototype fits seamlessly on both the prosthetic socket and pylon, demonstrating no misalignment during the assembly process and bench alignment (figure 8).

**Figure 8:**
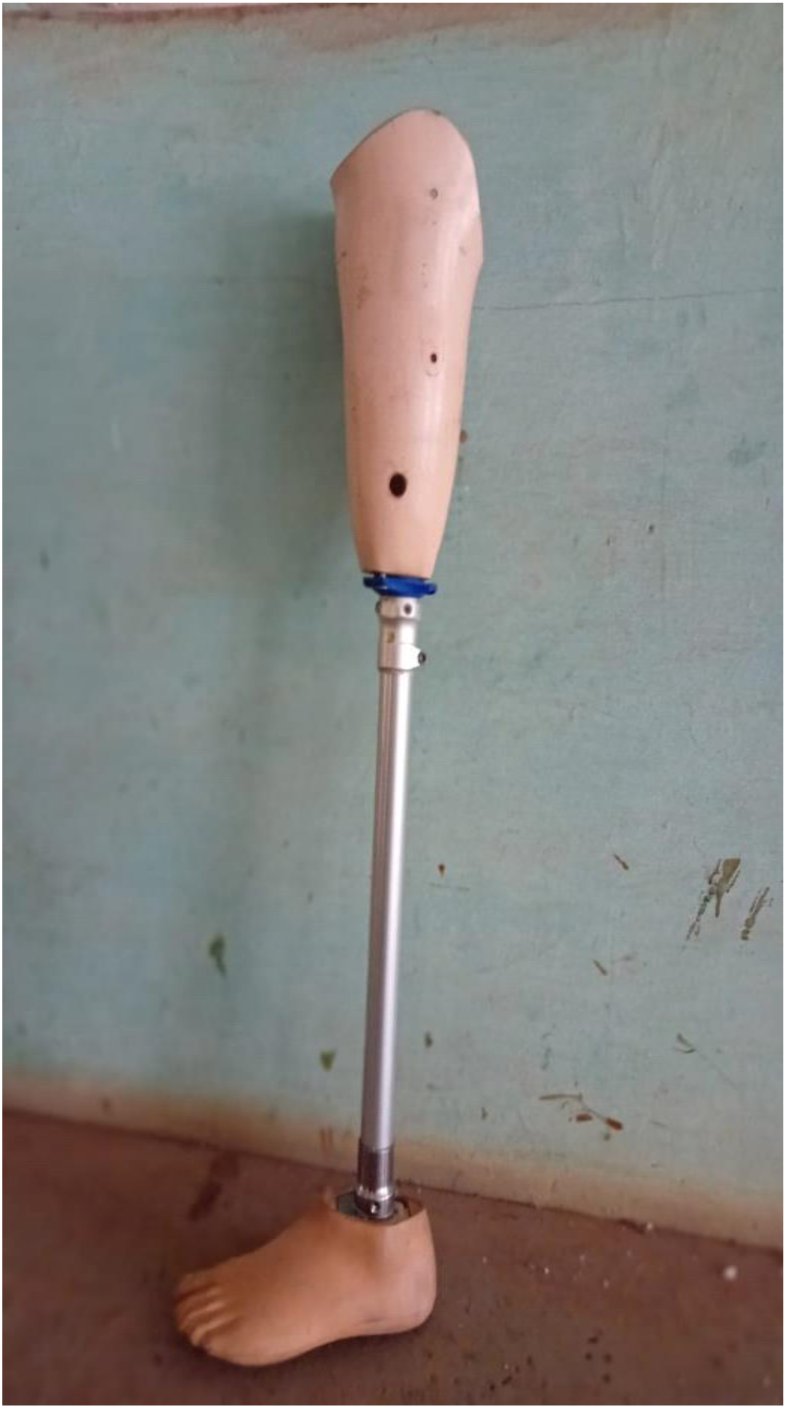
3D-printed socket adapter integrated within a complete transtibial prosthesis.

## Discussion

The primary objective of this study was to design, computationally analyze, and physically validate a 3D-printed prosthetic socket adapter as a cost-effective and accessible clinical simulation tool for Prosthetics and Orthotics (P&O) education in resource-limited environments. Our findings affirm the viability of this approach, demonstrating that a PLA-based FDM printed adapter can achieve the necessary dimensional accuracy, structural integrity, and functional compatibility for effective hands-on training. Specifically, computational analyses successfully identified critical stress and displacement regions, guiding design optimization, while physical testing confirmed the prototype’s precise fit with standard prosthetic components and its ability to withstand loads significantly exceeding those anticipated in a simulation context. This work provides a tangible solution to the acute scarcity of affordable training materials, promising to enhance the practical learning experience for aspiring prosthetists.

The successful fabrication of the prosthetic socket adapter, achieving dimensional deviations of less than 0.1 mm, highlights the precision attainable with accessible FDM 3D printing technology, even when operating at a relatively high print speed of 700 mm/s. This level of accuracy is critical for ensuring that the adapter can securely interface with existing, standardized prosthetic components, thus offering a realistic training experience. The mechanical testing, demonstrating a load-bearing capacity up to 600 N before significant deformation, far surpasses the forces typically encountered during non-ambulatory clinical simulation, which primarily involves alignment, assembly, and static fit assessments. This robust performance, coupled with the confirmed seamless fit on the prosthetic socket and pylon, suggests that the adapter is not merely a visual aid but a functional component capable of enduring the repetitive handling and minor stresses inherent in an educational setting. Furthermore, the overall smooth surface finish with minimal layer lines, achieved with minimal post-processing, ensures a professional appearance suitable for a training device.

The application of 3D printing in this context represents a significant step towards democratizing P&O education, particularly when compared to traditional manufacturing approaches. Conventional fabrication methods, including casting, CNC machining, and manual welding, are inherently time-consuming, labor-intensive, and necessitate substantial investment in expensive machinery, extensive workshop space, and specialized materials [19, 23, 24]. These factors contribute to the prohibitively high cost of imported prosthetic components, exacerbated by taxes and shipping delays in low-income nations like Ghana, rendering them largely inaccessible for extensive student training [5, 25, 26]. In stark contrast, our 3D printing approach leverages rapid, on-demand part production with minimal setup, requiring substantially less initial capital investment (a 3D printer can be up to ten times cheaper than CNC machining equipment) and a smaller footprint [21, 22]. This directly translates to lower production costs per unit, making hands-on training tools genuinely affordable and locally reproducible, thereby mitigating the reliance on costly imports and aligning directly with the sustainable development goals for educational access [27].

Beyond the immediate field of prosthetics, the utility of 3D printing for creating affordable educational tools resonates with findings in diverse engineering and medical disciplines. For instance, in mechanical engineering education, researchers have successfully utilized 3D printing to create low-cost models for teaching complex mechanisms, fluid dynamics principles, and material science concepts (e.g., studies on 3D-printed wind turbine models or gear train assemblies) [28–30]. These studies often emphasize how tangible, interactive models, fabricated on-demand at a fraction of the cost of commercial equivalents, significantly enhance student comprehension and engagement, especially when abstract concepts are involved. Similarly, our work leverages this principle by transforming theoretical P&O concepts into practical, hands-on experiences.

The implications of this research also extend to surgical training and medical simulation. Papers published in fields like orthopedic surgery or neurosurgery frequently highlight the development of 3D-printed anatomical models for pre-surgical planning and resident training [31–33]. These models, often customized from patient-specific imaging data, allow trainees to practice complex procedures in a low-stakes environment, improving dexterity and spatial reasoning [33, 34]. While our adapter is not patient-specific, the underlying principle of creating realistic, functional replicas for skill development is analogous. The affordability of PLA-based FDM printing, as demonstrated in our study, makes such simulation tools accessible even to institutions with limited budgets, a common challenge in healthcare education globally.

Furthermore, this work aligns with broader trends in global health and humanitarian engineering. Various initiatives have explored 3D printing for producing medical devices, assistive technologies, or spare parts in remote or underserved areas. For example, studies on 3D-printed assistive devices like specialized gripping tools or simple diagnostic components emphasize the importance of local fabrication using accessible materials to overcome supply chain issues and high import costs [35–38]. Also prosthetics 3D printing has been explored for conflict zone African countries [25]. Our focus on a prosthetic training component directly addresses the upstream problem: equipping local professionals with the practical skills needed to address the broader need for prosthetic care in their communities. The adoption of PLA, while having noted limitations regarding long-term durability in patient use, is a strategic choice for educational simulation due to its cost-effectiveness, ease of printing, and widespread availability, making the solution practical for resource-limited educational settings.

## Limitations

While the current prototype effectively serves its purpose as a clinical simulation tool, certain limitations warrant consideration for future work. The reliance on PLA, while cost-effective and easy to print, means the adapter is primarily suitable for educational demonstrations and not for long-term patient use due which would require materials with higher mechanical properties, greater heat resistance, and superior moisture stability. Future iterations could explore higher-performance polymers like PETG or even carbon fiber reinforced composites for enhanced durability, should the application extend beyond training. Furthermore, while the current mechanical testing confirmed strength for simulation, comprehensive cyclic loading tests would be beneficial if the adapter were to be adapted for prolonged, dynamic use. Future research could also involve direct feedback from P&O students and instructors in Ghana to further refine the design and integrate additional features that enhance the pedagogical value.

## Conclusion

This research successfully demonstrates the feasibility and substantial benefits of leveraging 3D printing technology to develop an affordable and functional prosthetic socket adapter specifically for clinical simulation and educational use in resource-limited environments. Through a meticulous design process, validated by Finite Element Analysis, we achieved a prototype exhibiting precise dimensional accuracy (deviations less than 0.1 mm), robust mechanical integrity (withstanding loads up to 600 N), and seamless functional compatibility with standard prosthetic components. These key findings collectively validate the adapter as a viable, cost-effective, and reproducible training tool.

The methodology presented in this study offers a compelling and replicable framework for addressing critical resource scarcity in P&O education, particularly in regions where traditional training materials are prohibitively expensive or inaccessible. Furthermore, the success of this approach extends its potential beyond prosthetics. The adoption and replication of this rapid, cost-effective, and accessible 3D printing methodology in other educational and practical fields is strongly encouraged, especially in low-resource settings, where the development of custom, functional, and affordable training aids or specialized tools can significantly enhance learning outcomes and practical capabilities. The methodology presented offers a replicable framework for similar educational material development, fostering self-sufficiency and innovation in global health education. This work ultimately champions a new paradigm for democratizing access to essential educational resources, fostering local innovation and self-sufficiency in global health and engineering disciplines.

## Data Availability

All data produced in the present study are available upon reasonable request to the authors

## CRediT authorship contribution statement

All authors reviewed and approved the final version of the manuscript. **Prince Oduro:** Writing – review & editing, Writing – original draft, Visualization, Software, Resources, Methodology, Investigation, Formal analysis, Data curation, Conceptualization. **Cecil Owusu Bempah:** Writing – review & editing, Writing – original draft, Resources, Investigation, Formal analysis, Conceptualization. **Sadat Osei-Wusu:** Writing – review & editing, Writing – original draft, Visualization, Software, Resources, Methodology, Investigation, Data curation, Conceptualization. **Joel Adjei Boateng:** Writing – review & editing, Writing – original draft, Visualization, Investigation. **Daniel Opoku-Gyamfi:** Writing – review & editing, Writing – original draft, Visualization, Software, Methodology. **Kannan Govindan:** Writing – review & editing, Validation, Supervision**. Napoleon Abiwu:** Writing – review & editing, Validation, Supervision.

## Ethical statement

The study was designed and conducted without involving human participants, patient data, or animal subjects. Therefore, no institutional review board (IRB) or ethics committee approval was required.

